# Divergence Between Clinician and Patient Perspectives on Polygenic Embryo Screening: A Qualitative Study

**DOI:** 10.1101/2023.10.12.23296961

**Authors:** Dorit Barlevy, Ilona Cenolli, Tiffany Campbell, Remy Furrer, Meghna Mukherjee, Kristin Kostick-Quenet, Shai Carmi, Todd Lencz, Gabriel Lazaro-Munoz, Stacey Pereira

**Affiliations:** Center for Medical Ethics and Health Policy, Baylor College of Medicine, Houston, TX; Center for Bioethics, Harvard Medical School, Boston, MA; Sociology Department, University of California Berkeley, Berkeley, CA; Braun School of Public Health and Community Medicine, The Hebrew University of Jerusalem, Jerusalem, Israel; Institute of Behavioral Science, The Feinstein Institutes for Medical Research, Northwell Health, Manhasset, NY; Departments of Psychiatry and Molecular Medicine, Zucker School of Medicine at Hofstra/Northwell, Hempstead, NY; Department of Psychiatry, Division of Research, The Zucker Hillside Hospital Division of Northwell Health, Glen Oaks, NY; Department of Psychiatry, Massachusetts General Hospital, Boston, MA

**Keywords:** polygenic embryo screening, polygenic preimplantation genetic testing, qualitative research

## Abstract

**Objective:** To explore and compare the perspectives of clinicians and patients on polygenic embryo screening.

**Design:** Qualitative.

**Subjects:** Fifty-three participants: 27 reproductive endocrinology and infertility specialists and 26 patients currently undergoing in vitro fertilization or had done so within the last five years.

**Main Outcome Measures:** Qualitative thematic analysis of interview transcripts.

**Results:** Both clinicians and patients often held favorable views of screening embryos for physical or psychiatric conditions, though clinicians tended to temper their positive attitudes with specific caveats. Clinicians also expressed negative views about screening embryos for traits more often than patients, who generally held more positive views. Most clinicians were either unwilling to discuss or offer polygenic embryo screening to patients or were willing to do so only under certain circumstances, while many patients expressed interest in polygenic embryo screening. Both sets of stakeholders envisioned multiple potential benefits or uses of polygenic embryo screening; the most common included selection and/or prioritization of embryos, receipt of more information about embryos, and preparation for the birth of a predisposed or “affected” child. Both sets of stakeholders also raised multiple potential, interrelated concerns about polygenic embryo screening. The most common concerns among both sets of stakeholders included the potential for different types of “biases” – most often in relation to selection of embryos with preferred genetic chances of traits –, the probabilistic nature of polygenic embryo screening that can complicate patient counseling and/or lead to excessive cycles of in vitro fertilization, and a lack of data from long-term prospective studies supporting the clinical use of polygenic embryo screening.

**Conclusion:** Despite patients’ interest in polygenic embryo screening, clinicians feel such screening is premature for clinical application. Though now embryos can be screened for their genetic chances of developing polygenic conditions and traits, many clinicians and patients maintain different attitudes depending on what is specifically screened, despite the blurry distinction between conditions and traits. Considerations raised by these stakeholders may help guide professional societies as they consider developing guidelines to navigate the uncertain terrain of polygenic embryo screening, which is already commercially available.

**Funding Statement:** This study was supported by the National Institutes of Health’s Human Genome Research Institute [R01HG011711].

**Disclosure Statement:** SC is a paid consultant at MyHeritage.

**Attestation Statement:**

- Data regarding any of the subjects in the study has not been previously published unless specified.
- Data will be made available to the editors of the journal for review or query upon request.

**Data Sharing Statement:** Appendices 1 and 2 will be available as supplemental materials upon publication. De-identified coded transcript excerpts will be made available upon reasonable request to the corresponding author.

**Capsule:** Clinician and patient perspectives on polygenic embryo screening both diverge and overlap, inviting greater reflection on concepts of condition severity and health for the development of professional guidelines.

## INTRODUCTION

Polygenic embryo screening (PES), also known as preimplantation genetic testing for polygenic diseases (PGT-P), differs from preimplantation genetic testing for aneuploidies (PGT-A) or monogenic conditions (PGT-M) in meaningful ways.^1^ Compared to PGT-A and PGT-M, PES has the capacity to screen for physical and psychiatric health conditions (e.g., diabetes, depression) as well as physical and cognitive traits (e.g., height, intelligence) (1). However, PES relies on inherently probabilistic polygenic risk scores, which are not yet standardized (2) and have limited portability to individuals of non-European ancestry (3). Further, PES may have limited utility in selection for traits (4), while the potential utility for reducing risk of disease is more complex (5–7). Nevertheless, PES has the potential to significantly alter the clinical experience of patients undergoing *in vitro fertilization* (IVF), and to expand IVF to populations that do not have fertility issues (8).

PES is currently available in the U.S. and other countries that do not regulate what kind of genetic testing may be conducted on embryos (9). Though a few professional organizations currently oppose the use of PES (10–13), uptake of PES will be determined largely by how clinicians, IVF patients, and the public view its potential utility, costs, and harms – at both individual and societal levels. To understand the potential future trajectory of PES, elaborate on the full range of considerations, both for and against the use of PES, and inform relevant guidelines that may be developed by professional societies, this qualitative study is the first to investigate the perspectives of US-based reproductive endocrinology and infertility specialists (REIs) and IVF patients regarding this new type of embryo screening.

## METHODS

We conducted semi-structured individual interviews over internet-secured video calls lasting between 29 and 86 minutes, with an average of 52 minutes for clinicians and 54 minutes for patients, which were audio-recorded and professionally transcribed.

Our multidisciplinary team (bioethicists, social scientists, and statistical geneticists) developed interview guides based on a literature review of PES, including its utility and ethical concerns. Interview questions investigated interest in, potential uses, and concerns about PES (Appendices 1 and 2). We piloted the interview guides with two clinicians and two patients and no substantive changes were necessary. Interviews included an explanation of PES with visualizations of mock embryo reports based on published examples from a commercial lab (14). Participants self-reported demographic information.

This study was approved by the Baylor College of Medicine Institutional Review Board (IRB), protocol H-49262 with a waiver of written documentation of consent. All participants provided verbal consent and received a $50 gift card.

### Recruitment

Between January and December 2022, we recruited clinicians and patients via convenience, random, and snowball sampling until we assessed that we reached “saturation” – the point at which subsequent data collection no longer generated novel insights (15).

Clinician participants were U.S.-based practicing REIs. For convenience sampling, we invited clinicians known to the research team. For random sampling, we conducted a search of U.S.-based members of the Society for Reproductive Endocrinology and Infertility (SREI) [https://www.socrei.org/sectiondirectory] which resulted in a list of 776 individuals. We used a random number generator to target 250 clinicians and made up to three attempts to invite them to participate.

Patient participants had to be currently undergoing IVF or had done so within the last 5 years. As part of convenience sampling, we invited IVF patients known to the research team. To obtain a random sample of IVF patients, we collated a list of 453 fertility clinics across the U.S. from a public fertility services webpage, which is no longer active [https://cofertility.com/get-to-know-us/], and invited them to post a flyer in their clinics. These flyers gave interested patients instructions for contacting the study team. Five fertility clinics posted the flyer (two each in the West and South, and one in the Midwest).

#### Analysis

Using thematic analysis (16), we developed a codebook based on deductive themes driven by the interview questions and unanticipated themes generated inductively from the interview data. Using Dedoose, a qualitative analysis program, at least two team members independently coded each transcript. Subsequently, one team member abstracted subthemes from coded excerpts, with a second team member reviewing the abstractions to confirm or discuss and reconcile subthemes (17). Because concerns were numerous and clustered, we grouped them thematically according to a third level of abstraction (17). Finally, for each code, one team member developed a memo, highlighting key subthemes and exemplary quotes, and a second team member reviewed for accuracy and comprehensiveness.

## RESULTS

See Figure 1 for recruitment details. In total, 27 of 235 invited REIs agreed to be interviewed, resulting in an enrollment rate of 11%. Because of our passive approach (i.e., clinic flyers) for patient recruitment, we are unable to report an enrollment rate for patients.

**Figure 1:**
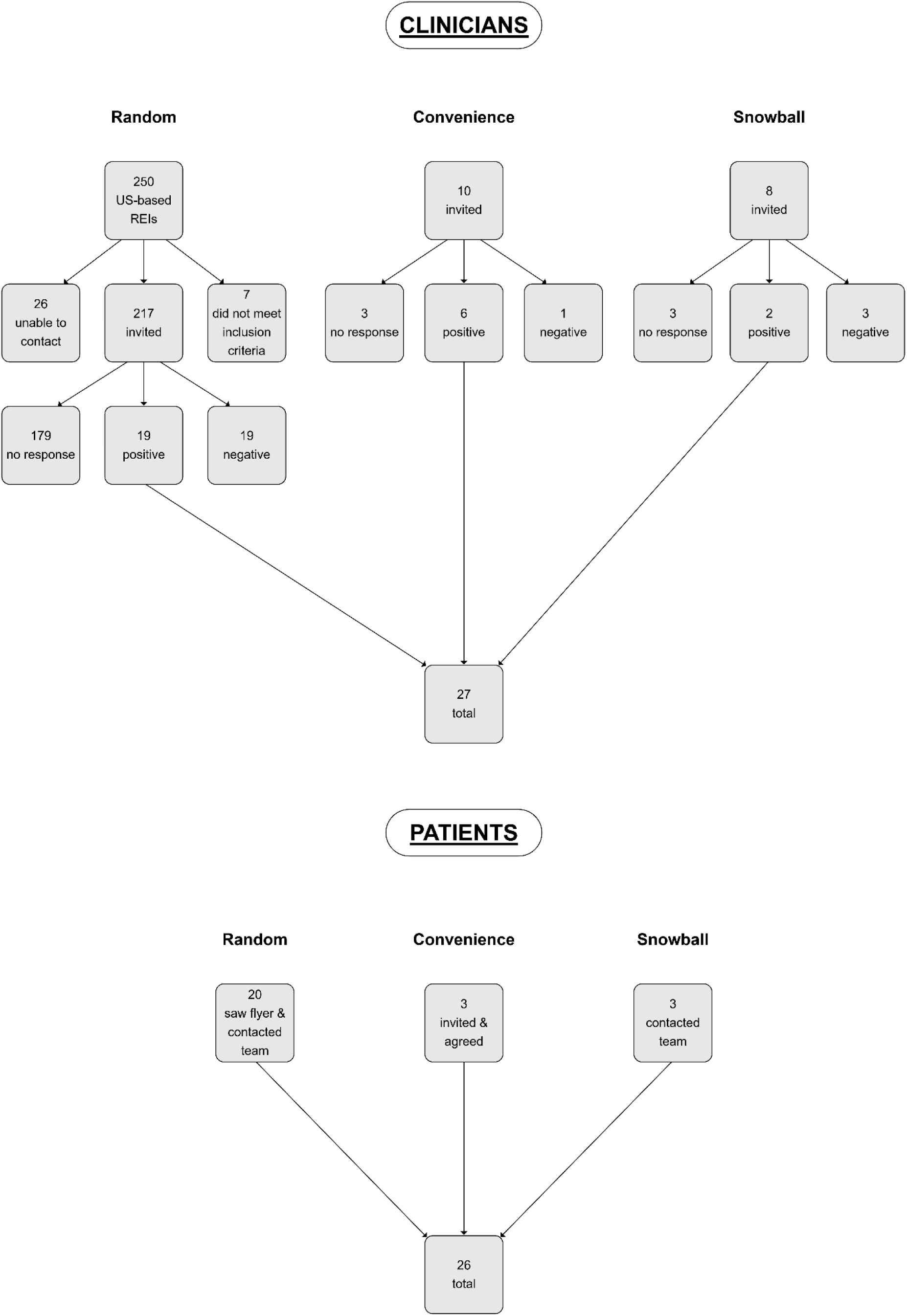
Recruitment Flow Chart.

Theoretical saturation was reached by interviewing 27 clinicians and 26 patients. Our sample of mostly White or European-American clinicians were situated across all U.S. regions, working in various practice types, and had a range of 3—40 years of REI experience, with a mean of 21.4 years. Patients were well-educated and wealthy, and most self-identified as female and White or European-American. Some had used or were planning to use preimplantation genetic testing. (For additional demographic details, see Tables 1 and 2.) Exemplary quotes and frequencies of subthemes are reported in Table 3.

**Table 1:** Clinician Demographics.

| Demographics |  | Counts | % of N=27 |
| --- | --- | --- | --- |
| Gender | Female | 12 | 44.4% |
|  | Male | 15 | 55.6% |
| Hispanic/Latino | Yes | 1 | 3.7% |
|  | No | 26 | 96.3% |
| Race* | American Indian, Native American, or Alaska Native | 0 | 0.0% |
|  | Asian | 5 | 18.5% |
|  | Black or African American | 0 | 0.0% |
|  | Native Hawaiian or Pacific Islander | 0 | 0.0% |
|  | White or European American | 20 | 74.1% |
|  | Other | 6 | 22.2% |
| Region | Northeast | 6 | 22.2% |
|  | Midwest | 6 | 22.2% |
|  | South | 9 | 33.3% |
|  | West | 5 | 18.5% |
|  | Other | 1 | 3.7% |
| Current Type of Practice | Academic | 8 | 29.6% |
|  | Military | 1 | 3.7% |
|  | Privademic | 5 | 18.5% |
|  | Private | 13 | 48.1% |

\*Respondents could report more than one category

**Table 2:** Patient Demographics.

| Demographics |  | Counts | % of N=26 |
| --- | --- | --- | --- |
| Gender | Female | 24 | 92.3% |
|  | Male | 2 | 7.7% |
| Hispanic/Latino | Yes | 4 | 15.4% |
|  | No | 22 | 84.6% |
| Race* | American Indian, Native American,<br>or Alaska Native | 0 | 0.0% |
|  | Asian | 7 | 26.9% |
|  | Black or African American | 0 | 0.0% |
|  | Native Hawaiian or Pacific Islander | 0 | 0.0% |
|  | White or European American | 18 | 69.2% |
|  | Other | 7 | 26.9% |
| Region | Northeast | 3 | 11.5% |
|  | Midwest | 3 | 11.5% |
|  | South | 6 | 23.1% |
|  | West | 14 | 53.8% |
|  | Other | 0 | 0.0% |
| Highest Level of<br>Education | Associate's degree | 2 | 7.7% |
|  | Bachelor's degree | 5 | 19.2% |
|  | Professional degree (JD) | 2 | 7.7% |
|  | Master's degree | 14 | 53.8% |
|  | Doctorate degree | 3 | 11.5% |
| Annual Household<br>Income Before<br>Taxes | \$75,000—\$99,999 | 3 | 11.5% |
| | \$100,000—\$149,999 | 7 | 26.9% |
| | \$150,000 or more | 16 | 61.5% |
| Number of IVF<br>Attempts | 1 | 14 | 53.8% |
|  | 2 | 11 | 42.3% |
|  | 3 or more | 1 | 3.8% |
| Current Number of<br>Children | 0 | 15 | 57.7% |
|  | 1 | 5 | 19.2% |
|  | 2 or more | 6 | 23.1% |
| Completed Family<br>Planning | Yes | 4 | 15.4% |
|  | No | 22 | 84.6% |

\*Respondents could report more than one category

**Table 3:**
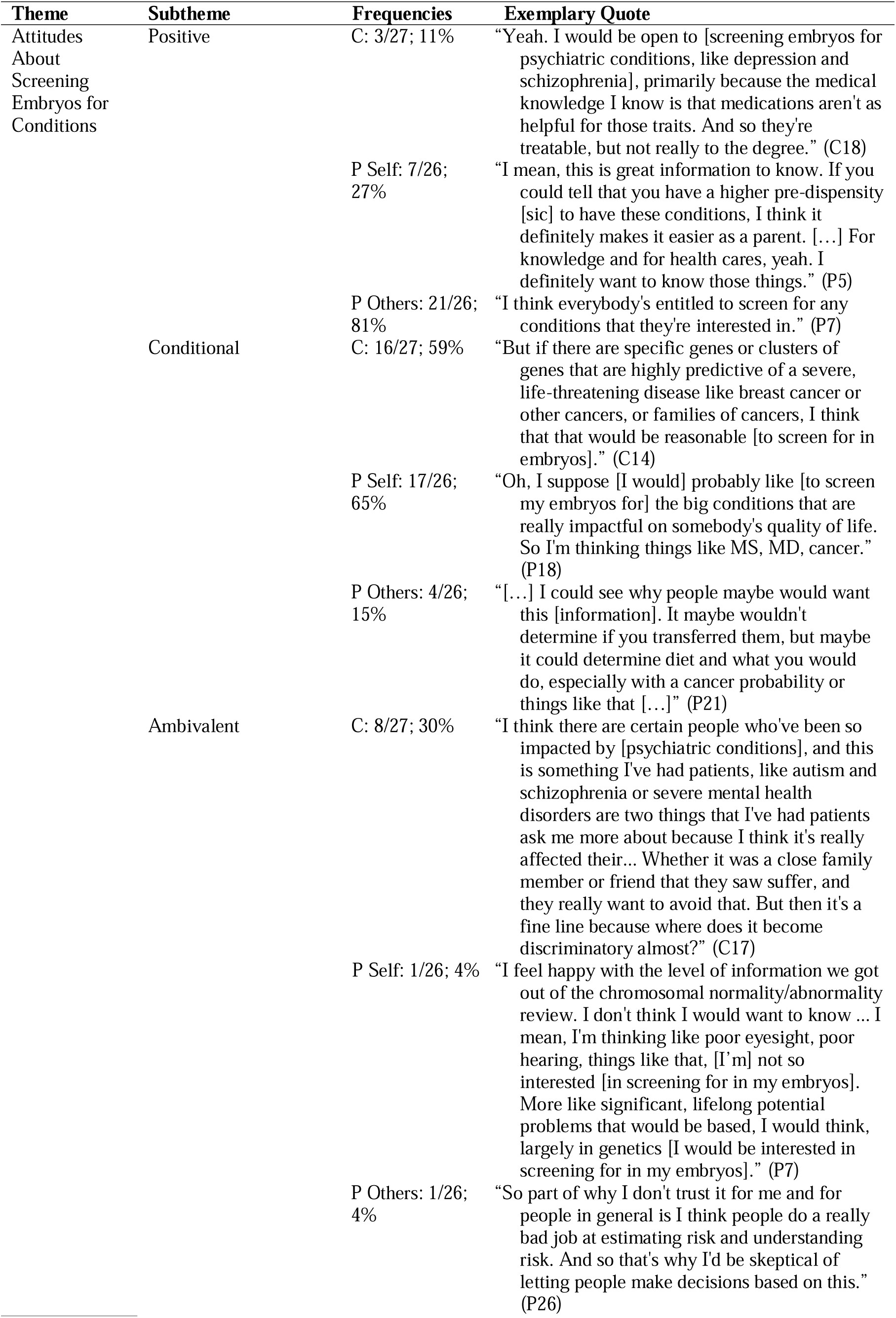
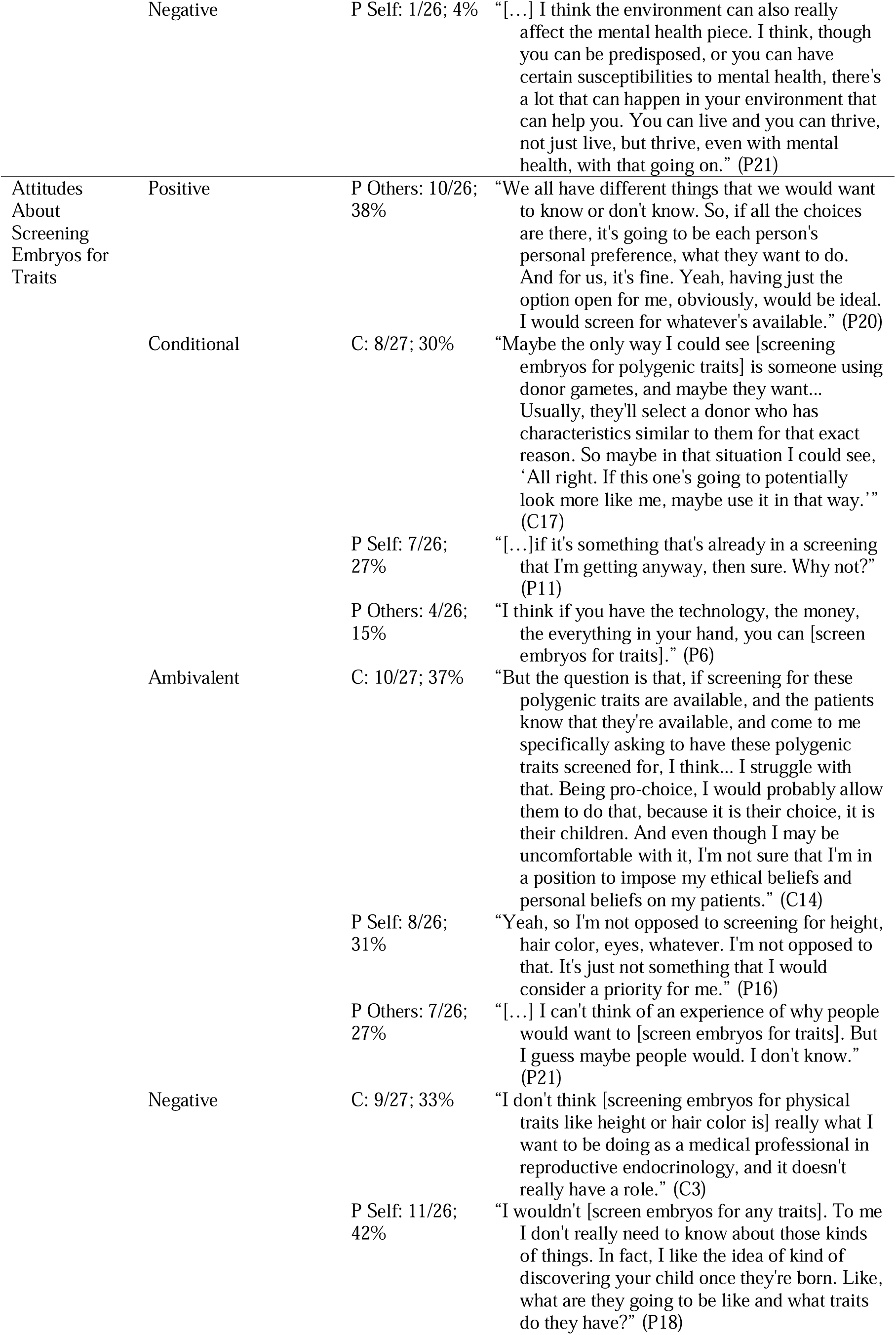
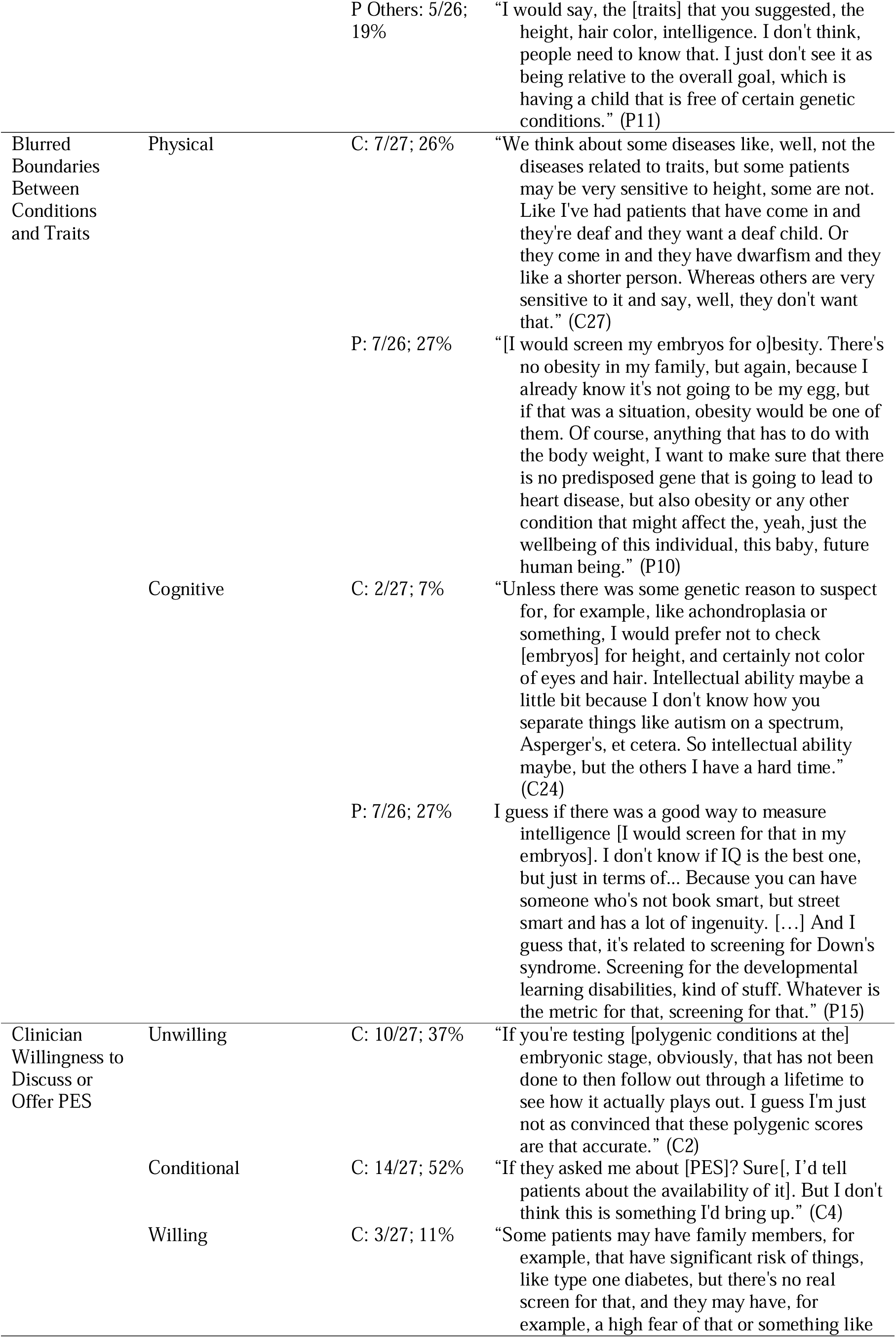
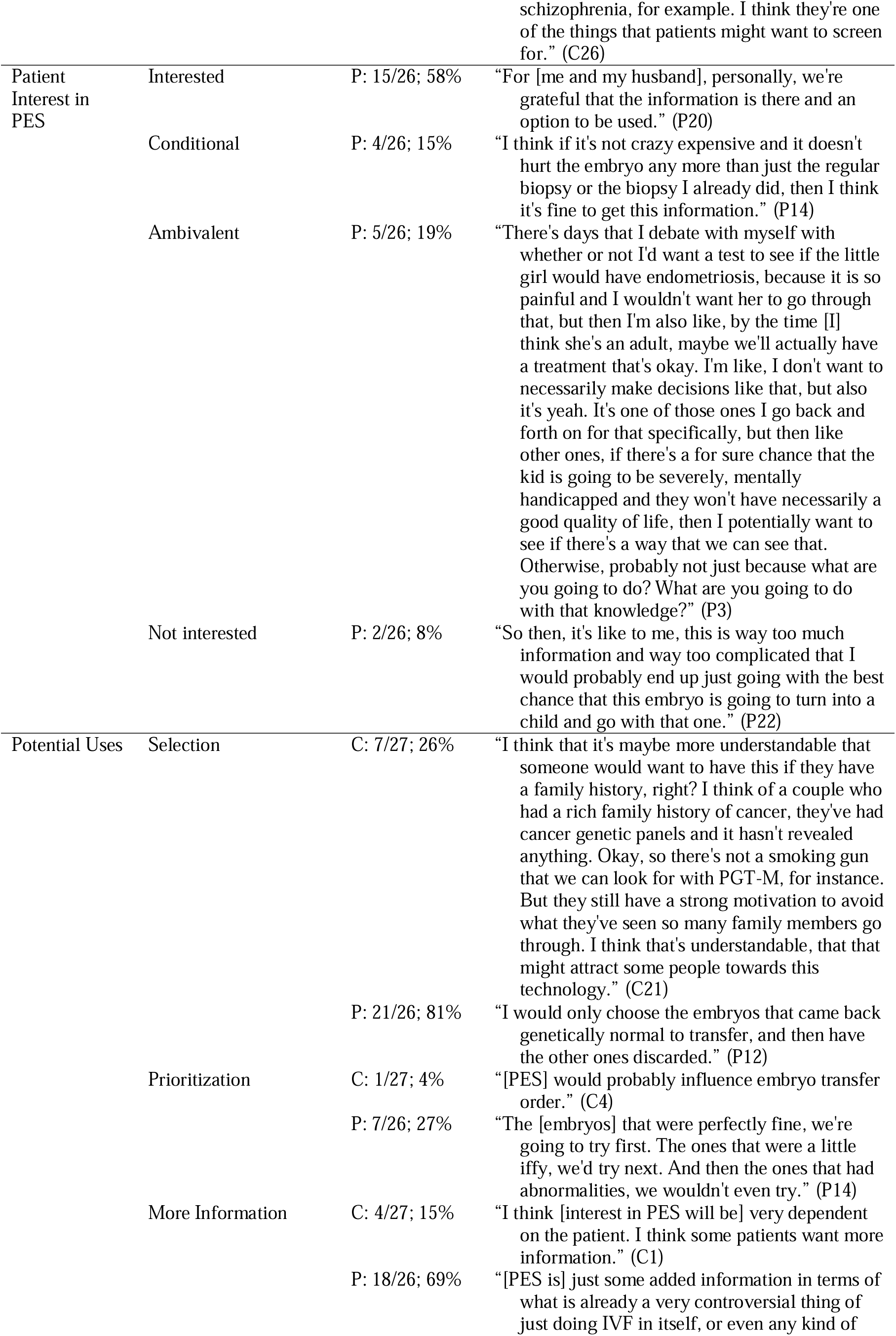
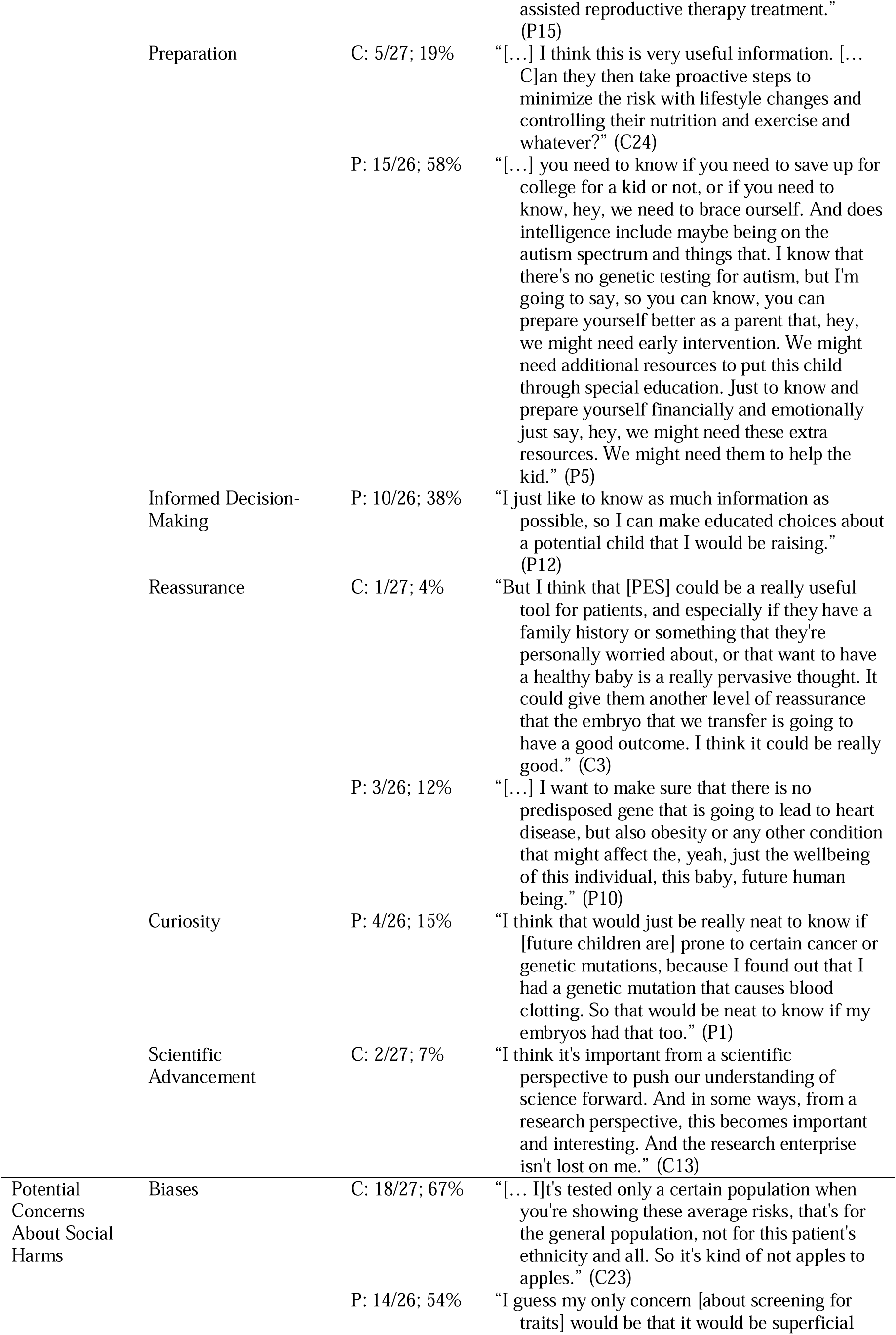
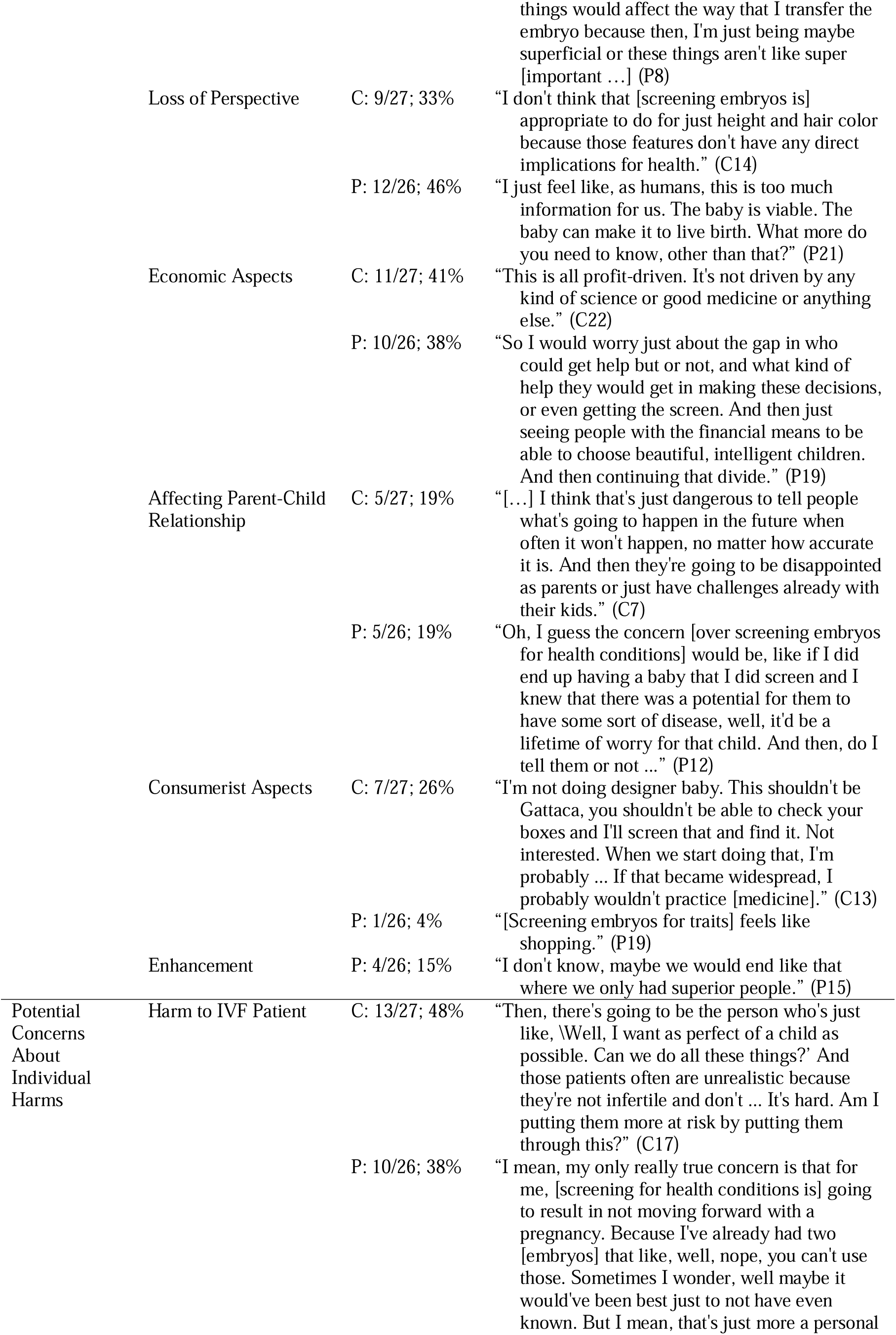
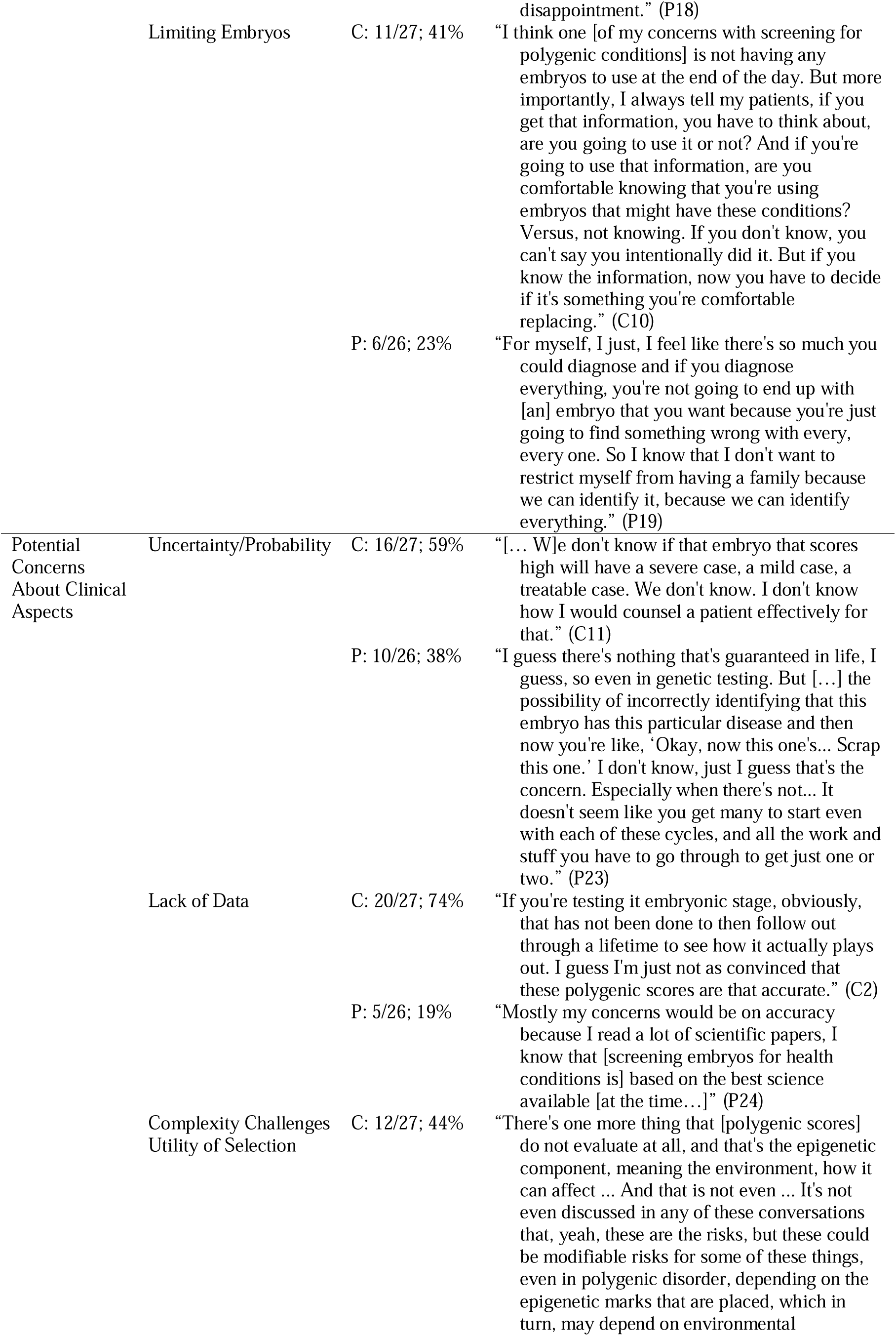
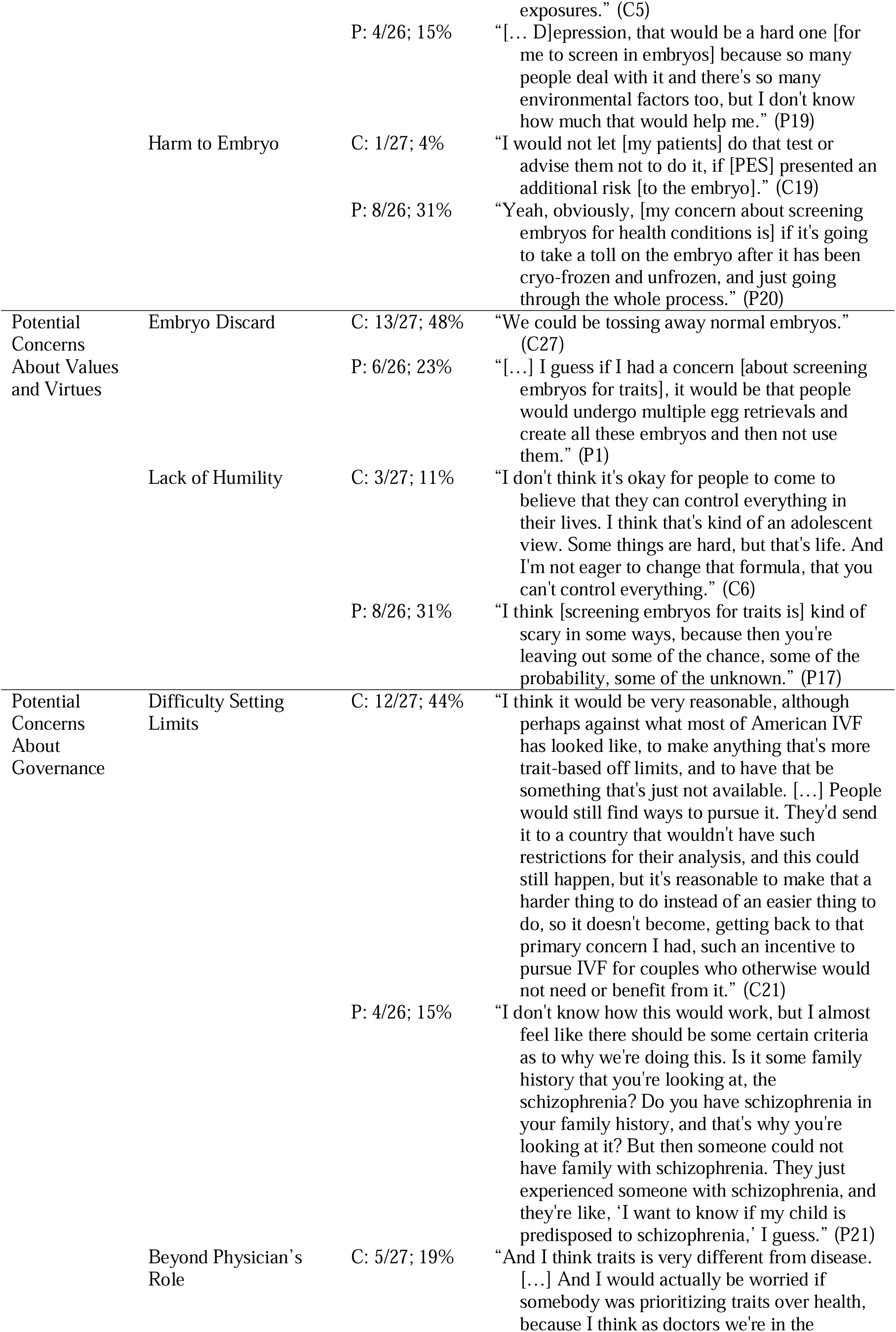
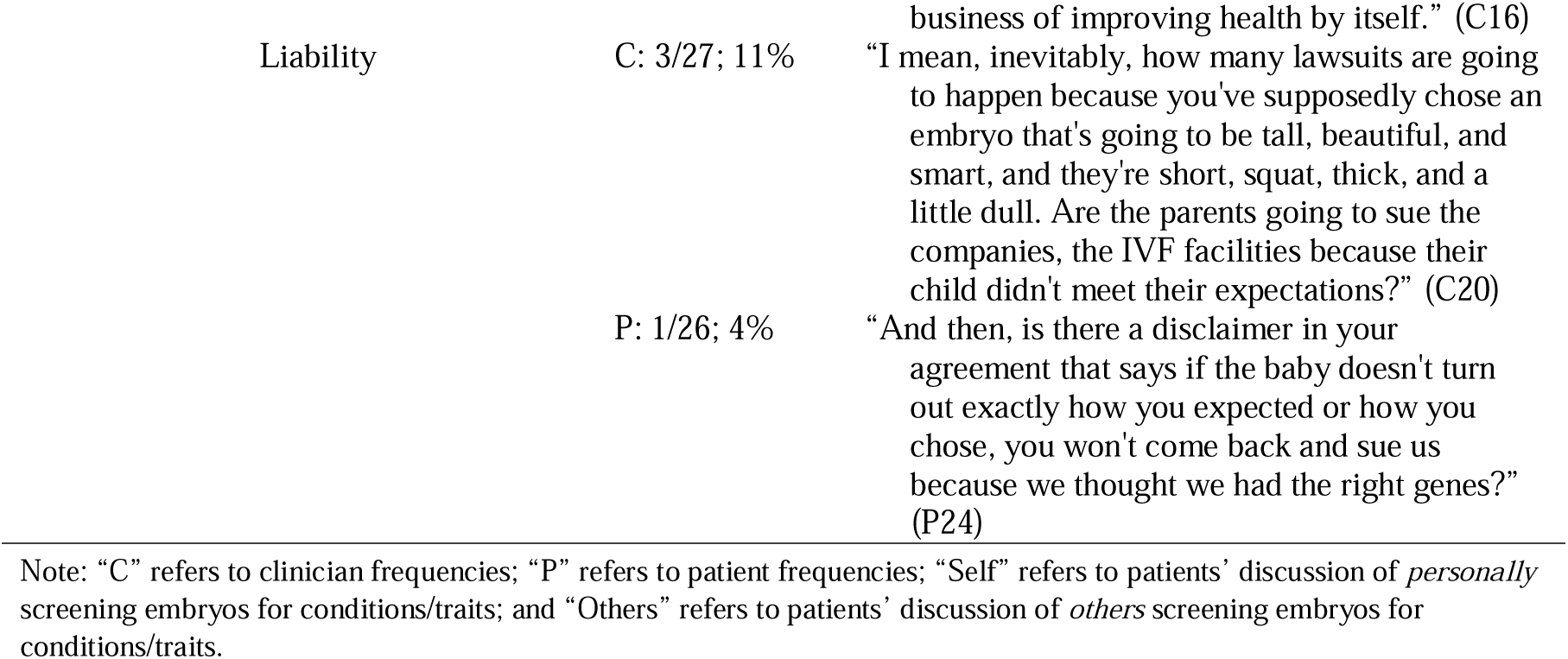
Exemplary Quotes.

### Attitudes about screening for conditions and/or traits

Clinicians and patients often held positive attitudes about screening embryos for physical and psychiatric conditions. However, clinicians tended to temper their positive attitudes with specific caveats, such as limiting embryo screening to conditions that are in one’s family history and/or considered severe. A minority of clinicians and patients opposed or felt ambivalent about screening for various conditions because of their variable severity, most often discussed with respect to psychiatric conditions (e.g., depression), or concern over how such information would be applied (e.g., discomfort with using it for selection). (For attitudes about specific conditions, see Figure 2.)

**Figure 2:**
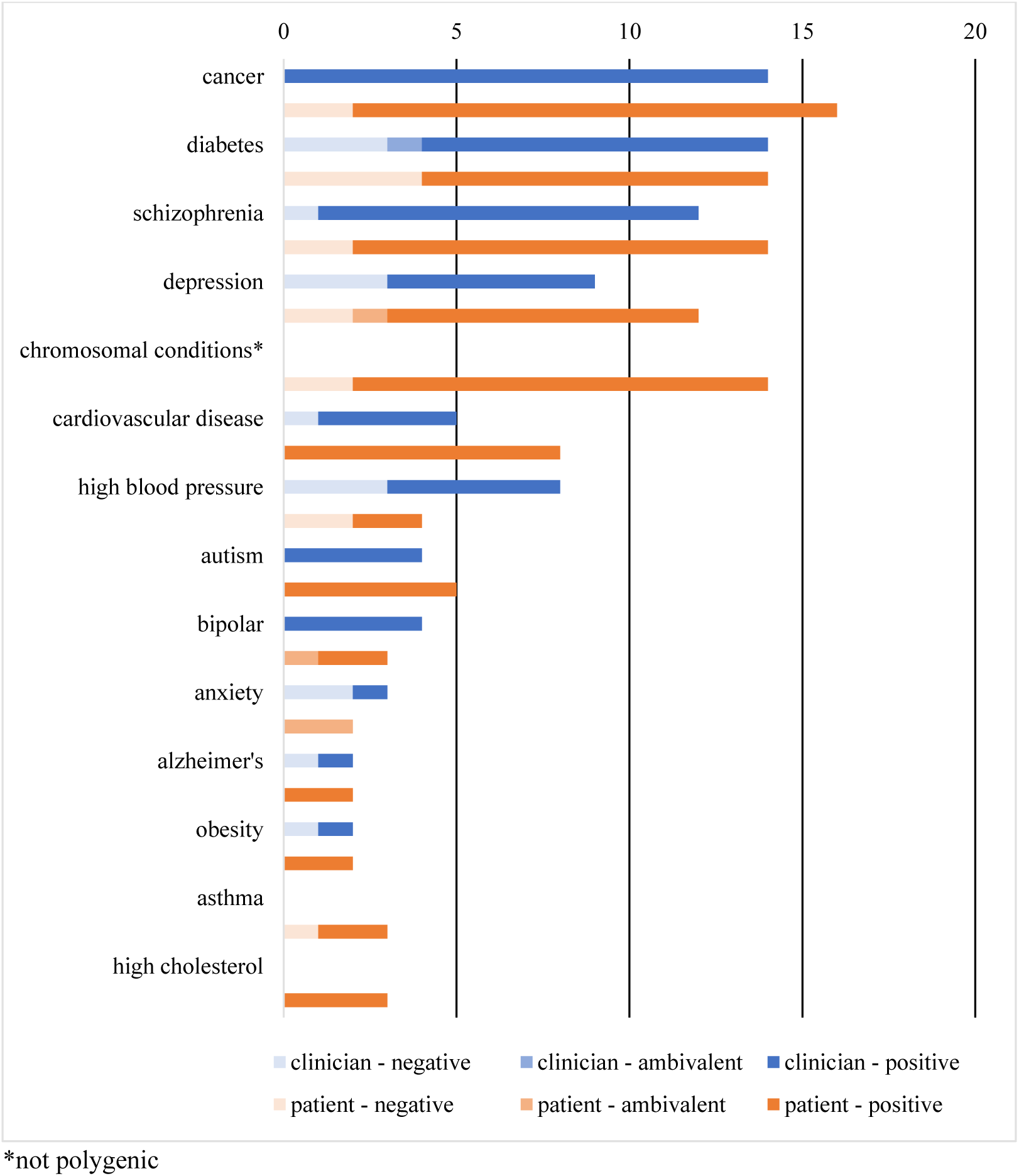
Attitudes on Screening Embryos for Most Discussed Conditions.

In contrast, screening embryos for traits generated greater variation of opinion. Clinicians expressed negative views about screening for traits more often than patients, who generally held more positive views. A minority of both sets of stakeholders were ambivalent about various traits. Intelligence was the most contentious trait; most clinicians that mentioned intelligence were against such screening, whereas most patients that mentioned it favored its screening. Opposition to screening embryos for traits was largely due to the belief that it is trivial, irrelevant to health or well-being, and/or beyond the role of medical professionals, as well as discomfort with using such information for embryo selection. Reasons for favoring screening for traits were largely based on respect for patients’ reproductive autonomy, even when some patients reported they were not interested in doing so themselves. (For attitudes about specific traits, see Figure 3.)

**Figure 3:**
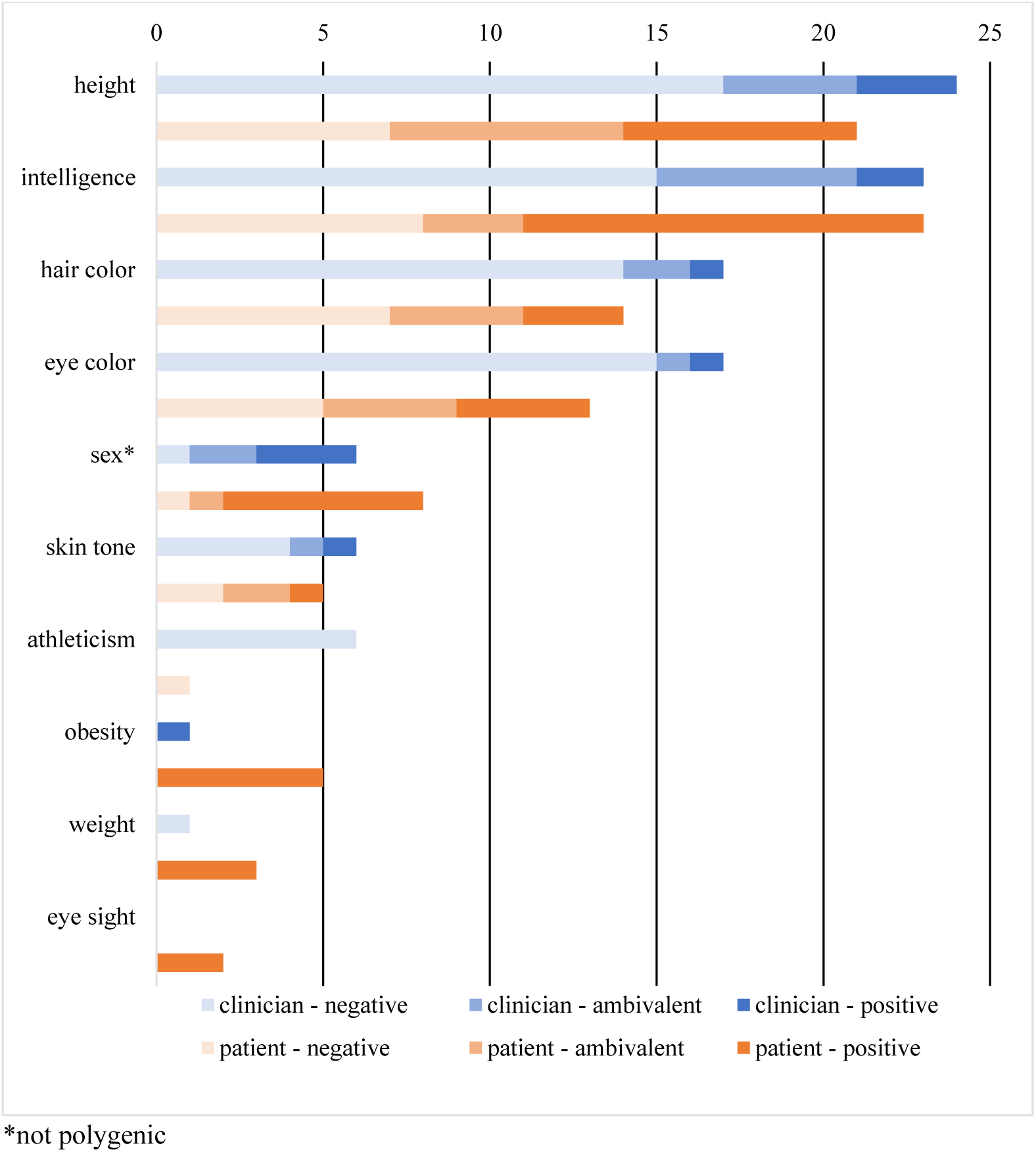
Attitudes on Screening Embryos for Most Discussed Traits.

Some clinicians and patients’ responses, however, suggested that the distinction between conditions and traits can be blurry. They occasionally mentioned obesity or achondroplasia when discussing screening for physical traits, and autism, Down syndrome, or learning disabilities when discussing screening for intelligence.

### Clinician reluctance to offer or discuss PES

Most clinicians were either unwilling to discuss or offer PES at this time or were willing to do so only under certain circumstances, most often if the topic was patient-initiated or PES was part of a research study. Some clinicians were willing to discuss or offer PES depending on the patient (e.g., those who, from the clinician’s perspective, were statistically savvy or had a compelling personal or family history of a polygenic condition), characteristics of the condition (e.g., untreatable), or external circumstances (e.g., more time for counseling; regulatory approval). Only a few clinicians were unconditionally willing to offer or discuss PES with patients at this time.

### Patient interest in PES

Despite clinicians’ hesitancy to offer or discuss PES, all 27 thought that at least some patients would be interested in such screening. Indeed, many patients expressed interest in PES during interviews. A few patients were interested in PES depending on various factors, such as financial costs or family history. Additionally, several patients maintained mixed feelings of interest (particularly for conditions they perceived as serious), tempered with multiple concerns (e.g., negative effects on parenting, potential for information overload, and boundaries around acceptable uses of PES). Two patients indicated they had no interest in PES, feeling it was too much information or irrelevant for their embryo selection.

### Potential uses of PES information

Though many clinicians perceived PES as potentially beneficial or useful for patients, some clinicians did not mention any potential benefits or uses of PES or said it was not beneficial or useful. In contrast, *all* 26 patients perceived PES as potentially beneficial or useful for themselves and/or other patients. Clinicians and patients who envisioned potential benefits of PES often mentioned multiple uses, including selection and/or prioritization of certain embryos; receipt of more information about one’s embryo(s); preparation for the birth of a predisposed or “affected” child; informed reproductive decision-making (i.e., using PES information to select embryos versus prepare for the birth of a certain child); reassurance of an embryo(s)’ lack of predisposition to certain conditions; satisfaction of curiosity; and scientific advancement when conducted for research. Clinicians and patients often portrayed embryo selection and prioritization as a means to: a) having a healthy (genetically-related) child, b) focusing on the “best” embryo, c) minimizing or preventing a future individual’s risk of developing a health condition, and/or d) improving society by reducing disease and/or creating “productive” members of society. Furthermore, clinicians and patients considered *selection and prioritization* most relevant in cases of multiple available embryos and/or predisposition for a condition(s) that was perceived as severe, often in terms of repeated or high morbidity, high/early mortality, compromising quality of life, lack of treatment, early onset, and/or affecting others. In contrast, clinicians and patients considered *preparation* most relevant in cases of few available embryos, regardless of perceived condition severity. Some clinicians and patients considered family history of a condition(s) to be either a reason or prerequisite for PES benefit or utility, especially in terms of justifying selection against certain embryos or preparation for the birth of a child with specific genetic risks.

### Potential concerns about PES

All 27 clinicians and 26 patients raised multiple potential, interrelated concerns about PES during the interview. Each set of thematically grouped concerns are listed in descending order of frequency across the entire sample of clinicians and patients. Notably, concerns over social harms were most numerous and frequent.

#### Social harms

The most common concern among all participants was the potential for different types of “biases.” Most often, this concern was in relation to embryo selection based on traits, with some clinicians and patients alluding to or specifically raising concerns about eugenics. Several clinicians and patients were concerned with political or subversive agendas, often referencing Nazi Germany, blue-eyed blonde Aryans, or creation of a “master race.” A few clinicians and patients worried that eugenic practices of selection following PES may (further) divide society or reduce human diversity. Additionally, some clinicians and patients raised concerns over bias inherent in the screening’s metrics (e.g., racial disparities in genome wide association studies) or the concept of measuring intelligence. Furthermore, a few patients and one clinician worried about potential physician bias in offering or counseling for PES; for example, offering it only to some patients or having personal perceptions influence how they counsel patients.

A common yet less frequent concern among clinicians and patients was the potential for loss of perspective as to what was important in life and/or IVF (e.g., valuing diversity and/or life itself, achieving pregnancy and live birth). Most of these concerns were made with respect to screening for traits, which was often considered trivial.

Another common yet less frequent concern among clinicians and patients was the economic aspects of PES. This concern was portrayed most often in terms of its added expense – which ultimately leads to unequal access to the technology – but also how the opportunity for profit drives its development.

Some clinicians and patients worried that knowledge about a selected embryo’s chances for developing a health condition or trait may negatively affect the parent-child relationship. Such negative effects may be due to resulting children either not living up to parental expectations or being treated as patients-in-waiting.

Finally, some participants raised concerns about PES’s consumerist aspects (e.g., designing babies; direct to consumer marketing) and the potential for PES to serve as a means for enhancement, in terms of creating “super” or “superior” people.

#### Individual harms

A common concern among clinicians and patients was the potential to harm IVF patients either psychologically or physically. Psychological harm was framed in terms of confusion, stress, or anxiety over PES information and what to do with it; disappointment if expectations are not met; and/or exploitation by companies offering PES. A few clinicians worried about PES’s potential to physically harm patients in cases when they electively seek to undergo IVF, with its associated risks, just to use PES, or undergo additional cycles of IVF to create or maximize embryo options. Such cases were portrayed as excessive.

Several patients, and even more clinicians, raised concerns about the potential for PES to limit or even eliminate embryos that patients perceive as acceptable for transfer. This concern reflected the use of PES for embryo selection in an IVF context where embryo availability is already limited.

#### Clinical and technical aspects

A common concern among clinicians and patients was over the uncertain or probabilistic nature of PES. Clinicians particularly worried that this would complicate counseling, especially amid time constraints, and/or lead to excessive IVF in pursuit of the “perfect” embryo. A few clinicians were unsure how to counsel patients about PES and worried about the lack of data and professional guidance on how to do so.

Clinicians’ most common concern, which several patients also shared, was a lack of data, usually with respect to the predictive value or generalizability of PES, because not enough research (e.g., long-term, prospective studies) has been conducted to support its clinical use. Some clinicians and patients were concerned specifically about a lack of data regarding antagonistic pleiotropy (i.e., genetic variants that lead to multiple phenotypes affecting evolutionary fitness in opposite ways) and/or more generally about the imperfect state of knowledge regarding genetics, human development, and health effects of IVF conception.

Some participants noted the multifactorial nature (i.e., interactions among genes, environment, and lifestyle) of polygenic conditions and traits as challenging the utility of PES for embryo selection. Often, this concern was made in association with screening embryos for cognitive traits (e.g., intelligence) and/or psychiatric conditions.

Some participants voiced general concerns about the potential of preimplantation genetic screening to physically harm the embryo(s). This was particularly acute for patients that experienced difficulties conceiving or those who ardently valued embryos’ potential for life.

#### Values and virtues

The concern of PES potentially leading to excessive or unethical embryo discard was common among clinicians but less so among patients. Most clinicians and patients with this concern specifically worried about discarding embryos characterized as “healthy,” “normal,” “fine,” or “viable.” Some clinicians and a few patients were concerned about discard resulting from a strive for perfectionism, which may lead patients to undergo excessive rounds of IVF.

Only a few clinicians but more patients felt PES represented a lack of humility in terms of accepting limitations to human control and/or knowledge. Though this concern was most often made with respect to screening (and selection) for traits, it was sometimes made regarding PES in general or screening for (and selection against) health conditions.

#### Parameters and governance

A rather common concern among clinicians, but not as much for patients, was difficulty in setting limits as to what is acceptable to screen for in embryos and who should be permitted to use PES. Some clinicians and patients felt that screening embryos to select against those with increased genetic risk for manageable or treatable conditions was inappropriate because individuals at increased risk or even with the condition can lead fulfilling and healthy lives. Additionally, some felt that screening embryos to select against those with (increased genetic risk for) conditions with: a) adult onset is problematic because of the decades required for studies to validate the data and the potential for medicine to develop treatments by the time of onset, or b) low (absolute or relative) risk may not be worthwhile, considering the potential to modify such risk(s) via environment or lifestyle. Furthermore, several clinicians and patients noted the ever-changing contextual nature of classifying some traits as desirable and selecting embryos based on their likelihood for developing them.

Several clinicians felt embryo selection based on PES, particularly for traits, was not part of a physician’s role, which is focused on treating disease (i.e., infertility in the case of REIs). Hence, facilitating such selection would be beyond their medical scope.

A few participants worried about potential liability issues when selected embryos do not meet IVF patients’ expectations for their eventual children (e.g., of developing certain traits or not developing certain conditions).

## DISCUSSION

This study, which is the first to compare clinician and patient perspectives of PES, yielded several noteworthy findings. First, there appears to be a gap between clinician and patient attitudes toward PES, whereby clinicians generally maintained reservations about such screening and patients indicated interest in it. This finding aligns with recent studies of American and European healthcare professionals’ attitudes toward PES (18,19), American IVF patients’ increasing use of preimplantation genetic testing (20), high acceptance of PES (21), and high uptake of PES when offered at no additional financial cost to patients that used PGT-A (22). Moreover, REIs’ greatest concern about the lack of available data to support PES may reflect their perceptions of the controversial widespread clinical implementation of PGT-A, which some have argued was premature (23,24).

Interestingly, though PES is marketed and usually discussed as a tool for embryo selection (7), we found that clinicians and patients sometimes envisioned PES being used to prepare for the birth of a predisposed or “affected” individual. Although the intentional or incidental transfer of embryos with pathogenic variants detected in preimplantation testing is rare (25), preparation has not been reported previously as a motivation or decisional factor for using preimplantation genetic testing (26). However, preparation has been considered a benefit or use of *prenatal* testing (27). Thus, a conflation between prenatal and preimplantation genetic testing may be a factor contributing to these comments. Further research is warranted to determine whether PES’s potential for preparation reflects a real or theoretical use of the screening information.

Another notable finding is the difference in opinion between screening embryos for conditions versus traits among both clinicians and patients, with far less support for traits. This finding aligns with previous measured stances of ASRM’s Ethics Committee regarding embryo sex disclosure and selection (28,29) and previous studies of pregnant women’s perspectives of noninvasive prenatal testing and whole genome sequencing (30,31). Yet some clinicians’ and patients’ references to certain *conditions* while discussing screening for *traits* blurred the distinction between these two categories. Perhaps it may be more apt to consider PES in terms of screening embryos for health related- and non-health related-traits, especially when deliberating on the potential guidance for it.

Relatedly, severity and definitions of health loom large in discussions of PES. Previous studies report condition severity is a main factor in deciding whether to use preimplantation genetic testing, and the ability to control or improve the health of one’s future child(ren) is a main motivation for using it (26,32). However, the constitution of severity and health is debatable (33–36). Though some scholars believe consensus on defining these terms is impossible (37), others propose developing an adaptable framework that incorporates biomedical, social, and personal meanings (38). Either way, the prospect of PES invites clinicians, IVF patients, and all of society to contemplate the meanings of these concepts.

### Limitations

This qualitative study’s findings may be limited by social desirability response bias (i.e., answering questions in a manner that will be viewed favorably by others) and self-selection bias. Thus, its generalizability to other REIs and IVF patients may be limited. Furthermore, discussion of screening embryos for specific conditions and traits sometimes was prompted by the interviewer: a) citing examples of heart disease, cancer, depression, schizophrenia, height, hair color, and intelligence, and b) demonstrating a PES report, which included conditions such as diabetes and cancer (Appendices 1 and 2).

## CONCLUSION

Despite patients’ interest in PES, clinicians feel such screening is premature for clinical application. Though now embryos can be screened for their genetic chances of developing polygenic conditions and traits, many clinicians and patients maintain different attitudes depending on what is specifically screened, even though the distinction between conditions and traits is not always clear-cut. This dual-use aspect will prove challenging for governing PES. Professional societies are best positioned to develop guidelines for navigating the uncertain terrain of PES, which is already commercially available. Moreover, there should be greater discussion within medicine and society over meanings of “severe” and “health.”

## Data Availability

De-identified coded transcript excerpts will be made available upon reasonable request to the corresponding author.

## ACKNOWLEDGEMENTS

The authors thank the REIs and IVF patients that participated in this study, as well as Jason Bach, Arturo Balaguer, Jonathan Frumovitz, and Page Trotter for their assistance with data analysis and Ana Battaglino for her feedback on the manuscript.

## Footnotes

1 We use the acronym PES (vs. PGT-P) to highlight these differences from other types of preimplantation genetic tests.

## Notes

### Funding Statement

This study was funded by the National Institutes of Health, Human Genome Research Institute [R01HG011711]

### Author Declarations

The IRB of Baylor College of Medicine gave ethical approval for this work.

